# Mapping the costs of mental health- and substance use-related grant cancellations

**DOI:** 10.1101/2025.08.01.25332818

**Authors:** Ariel L. Beccia, Lingbo Liu, Scott Delaney, Dougie Zubizarreta, Noam Ross, S. Bryn Austin

## Abstract

**Background:** Since the 2025 Presidential Inauguration, the Trump Administration has terminated billions of dollars in federal funding for science; however, the impacts of these grant terminations on the mental health and substance use fields have not yet been examined. We thus aimed to quantify and map the costs associated with federally funded mental health- and substance use-related grants that have been prematurely terminated.

**Methods:** We used a comprehensive dataset of grants terminated by the National Institutes of Health (NIH), National Science Foundation (NSF), and Substance Abuse and Mental Health Services Administration (SAMHSA) compiled from multiple sources. After identifying terminated mental health- and substance use-related grants from this database via a two-step screening process, we quantified their number and associated lost funding for each congressional district, which we visualized using a series of maps to examine trends and regional variations.

**Outcomes:** We identified 474 mental health- and/or substance use-related grants that were terminated by the NIH, NSF, or SAMHSA from February 28, 2025, through April 11, 2025, totaling $2,098,731,548 in lost funds. Congressional districts corresponding to urban centers with large academic and research institutions (e.g., New York City, Boston) experienced the most pronounced losses from NIH and NSF grants, whereas districts located throughout the Mid-Atlantic, Midwest, Southeast, and Southwest were the hardest hit by the termination of SAMHSA block grants (i.e., those used to pay for community mental health and substance use services).

**Interpretation:** Against a backdrop of ongoing and intersecting mental health and substance use crises, the Trump Administration has slashed research dollars on these topics, creating a chilling effect on the field. Such cuts are likely to destabilize existing mental health and substance use services and exacerbate inequities between and within U.S. states, ultimately intensifying the challenges faced by local communities.

**Funding:** None to report.

## Introduction

Almost immediately following Donald Trump’s inauguration as the 47^th^ president of the U.S., he signed a series of executive orders that would prove to hold disastrous implications for medical and public health research. Most pertinently, Executive Order 14151 sought to terminate all government programs related to “diversity, equity, and inclusion” (i.e., DEI),^1^ while Executive Order 14168 declared sex to be “an individual’s immutable biological classification as either male or female” and demanded an end to any federal recognition of trans and nonbinary identities;^2^ both significantly narrowed the scope of permissible topics and populations that federally funded research could focus on or even mention, and introduced particular challenges to conducting equity-focused work. Indeed, and in conjunction with the broader right-wing ideologies of the Trump Administration (e.g., those characterized by conservatism, nationalism, and market fundamentalism), these orders and related policies have since been used as justification for the executive branch to interfere with the conduct of biomedical and social science in the U.S. so as to preclude scientific inquiry into areas deemed “irrelevant,”^3^ including terminating grants awarded to local health departments, non-profit organizations, and academic institutions that were focused on (or were perceived to be focused on) “DEI” or sex/gender, as well as the COVID-19 pandemic, global health, or climate change.^4^ Since the inauguration, billions of dollars in federally funded grants have been terminated, with increasing reports highlighting how those on mental health and substance use have not been spared.^5^ Such actions would stand in stark contrast to the Trump Administration’s professed support for addressing our ongoing mental health and substance use (e.g., opioid) crises and for “making America healthy again”^6^ and are also expressly concerning given the escalating rates of and widening inequities in depression, suicide, and addiction amongst Americans.^7,8^ Moreover, the lost funds are likely to have devastating impacts on local economies, likely reducing critical resources and exacerbating challenges for affected communities.^9^

While several reports have characterized the extent and breadth of grant terminations by government agencies and across geographies, most have been general in their examination or have focused on targeted topics and populations (e.g., gender affirming care, LGBTQ+ people);^10–12^ as such, the impacts of terminations on the fields of mental health and substance use research more broadly remain unknown. We thus aimed to address this gap by quantifying and mapping the costs associated with federally funded mental health- and substance use-related grants that were prematurely terminated by the Trump Administration from Inauguration Day, January 20, 2025, through April 11, 2025. To provide further context, we also aimed to characterize canceled grants by funding agency or institute, mechanism, and topic area. Informed by and building upon recent analyses published in *JAMA, Nature*, and *STAT*,^10,11,13^ as well as the revelatory U.S. Senate Committee on Health, Education, Labor, and Pensions (HELP) “Trump’s War on Science” report,^12^ we mapped terminations by congressional district, a unit of political geography that is highly relevant to policymakers and community advocates.

## Methods

We used a publicly available dataset of grants terminated by the National Institutes of Health (NIH), National Science Foundation (NSF), and Substance Abuse and Mental Health Services Administration (SAMHSA) compiled from several sources, including affected principal investigators, who submitted details of their terminated grants using a Google Form that was designed and published specifically for this purpose.^14^ The dataset also aggregated data from news reports, social media, Doge.gov, USASpending.gov, NIH’s X feed, NIH RePORTER, the Department of Health and Human Services’ (HHS’) Tracking Accountability in Government Grants System (TAGGS) of grant-based financial transactions, and a publicly available, HHS-generated PDF list of terminated grants. Because we used a dataset that aggregated data from these multiple sources, it included grants not listed on federal government websites, and thus comprised the most comprehensive, up-to-date resource available that quantified and characterized terminated grants.

To identify mental health- and substance use-related grants from this database, we implemented a multi-step procedure. First, based on our team’s content-area expertise, we developed a preliminary search strategy composed of keywords likely to be included in relevant grants. Second, we conducted several rounds of iterative co-author feedback and testing to ensure the keywords were comprehensive and specific. Third, once we derived the final search strategy (presented in the Supplement), we developed a Python-based pipeline to identify relevant grants using keyword matching in their project summaries, project narratives, terms, and a column of derived relevant “flagged words” (i.e., those identified by *The New York Times* as words the federal government sought to purge from federal websites because they related to topics the government deemed unfavorable^15^). Fourth, the resultant list of relevant grants was screened by two authors (ALB and DZ) to assess whether they met our eligibility criteria (i.e., directly examining, analyzing, or addressing topics related to mental health and/or substance use in their project aims or objectives). After excluding 108 of 582 identified grants, we had a final list of 474 that were terminated during the period from February 28, 2025 (the date of the first wave of terminations) through April 11, 2025 (the end date in our dataset).

Subsequently, to analyze the impact of these terminations, we quantified their number and associated lost funding for each congressional district. For grants lacking geographic information, we performed geographic decoding by cross-referencing state and recipient organization names with congressional district boundary data. The resulting records were aggregated by congressional district, allowing us to calculate both the total funds originally awarded and the remaining funds that were lost due to premature termination. We then visualized lost funding on a congressional district-level map to examine trends and regional inequities in mental health and substance use funding forfeiture across the U.S. As mentioned, we also examined descriptive information on terminated grants’ original funding agency or institute, mechanism (e.g., R-series), and topic area.

## Results

We identified 474 mental health- and/or substance use-related grants that were terminated from February 28, 2025, through April 25, 2025, totaling $2,098,731,548 in lost funds. Most terminated grants were from the NIH (n=243, 51.2% of grants) or SAMHSA (n=224, 47.3%); only seven (1.5%) relevant NSF grants had been terminated by April 25. Table 1 presents additional descriptives for terminated NIH grants, which were highly diverse in terms of focus; conversely, 214 of the 224 terminated SAMHSA grants were block grants awarded to state health departments and the remaining 10 were emergency COVID-19 pandemic funds. Among terminated NIH grants, many were funded by the National Institute of Mental Health (NIMH; n=69, 28.4%), followed by the National Institute on Drug Abuse (NIDA; n=46, 18.9%) and the National Institute on Alcohol Abuse and Alcoholism (NIAAA; n=33, 13.6%). The majority were R-series grants (n=172, 70.8%), driven by the large number of terminated R01s (105 of 172), although there were a considerable number of training grants (i.e., F-, K-, and T-series) grants that were terminated as well (n=48, 19.8%). Regarding topic areas, over half of the terminated NIH grants were for projects focused on LGBTQ health (n=174, 71.6%) or race/racism (n=169, 69.5%).

**Table 1.** Characteristics of grants terminated by the National Institutes of Health (NIH) (N=243)

|  | **Terminated grants**<br>**(N = 243)** |
| --- | --- |
| **Total lost funds, in dollars** | 710,800,838 |
| **Funding agency or institute, No. (%)** |  |
| Fogarty International Center (FIC) | 2 (0.8) |
| National Cancer Institute (NCI) | 7 (2.9) |
| National Heart, Lung, and Blood Institute (NHLBI) | 5 (2.1) |
| National Human Genome Research Institute (NHGRI) | 1 (0.4) |
| National Institute of Allergy and Infectious Diseases (NIAID) | 4 (1.6) |
| National Institute of Arthritis and Musculoskeletal and Skin Diseases (NIAMS) | 1 (0.4) |
| National Institute of Biomedical Imaging and Bioengineering (NIBIB) | 1 (0.4) |
| National Institute of Child Health and Human Development (NICHD) | 20 (8.2) |
| National Institute of Environmental Health Sciences (NIEHS) | 3 (1.2) |
| National Institute of General Medical Sciences (NIGMS) | 2 (0.8) |
| National Institute of Mental Health (NIMH) | 69 (28.4) |
| National Institute of Minority Health and Health Disparities (NIMHD) | 28 (11.5) |
| National Institute of Neurological Disorders and Stroke (NINDS) | 2 (0.8) |
| National Institute of Nursing Research (NINR) | 9 (3.7) |
| National Institute on Aging (NIA) | 9 (3.7) |
| National Institute on Alcohol Abuse and Alcoholism (NIAAA) | 33 (13.6) |
| National Institute on Drug Abuse (NIDA) | 46 (18.9) |
| NIH Office of the Director (OD) | 1 (0.4) |
| **Grant mechanism, No. (%)** |  |
| D-series (D43, DP2, DP5) | 4 (1.6) |
| F-series (F30, F31, F32) | 25 (10.3) |
| K-series (K01, K08, K12, K32, K24, K99) | 14 (5.8) |
| O-series (OT2) | 1 (0.4) |
| P-series (P01, P50) | 2 (0.8) |
| R-series (e.g., R00, R01, R03, R21, R24, R25, R33, R34, R36, R37, R41) | 172 (70.8) |
| S-series (S21, SC1, SC3) | 3 (1.2) |
| T-series (T32) | 9 (3.7) |
| U-series (U01, U54, UG3, UM1) | 13 (5.3) |
| **Topic area (non-mutually exclusive), No. (%)** |  |
| Transgender health-related | 72 (29.6) |
| LGBTQ+ health-related | 174 (71.6) |
| HIV-related | 66 (27.2) |
| Race or racism-related | 169 (69.5) |
| Vaccine-related | 7 (2.9) |

*Note:* Percentages may not sum to 100% due to rounding. Additional descriptives are not provided for grants terminated by the National Science Foundation (NSF) or the Substance Abuse and Mental Health Services Administration (SAMHSA), as these were fewer in number and less diverse in focus.

Figure 1 presents the lost funding due to NIH and NSF (1a) and SAMHSA (1b) grant terminations by congressional district; the labels highlight the 10 districts that lost the greatest amount of funds. Focusing first on NIH and NSF grants (combined due to their near-exclusive focus on research and/or training), we found that North Carolina’s 4^th^ congressional district lost the most funding due to terminated mental health- and substance use-related grants at $478 million, followed by New York’s 13^th^ district ($50 million) and Virginia’s 5^th^ district ($24 million). In general, congressional districts corresponding to urban centers with large academic and research institutions, including Durham (e.g., Duke University, UNC-Chapel Hill), New York City (e.g., Columbia University), and Boston (e.g., Harvard University, MIT), experienced the most pronounced losses. In terms of SAMHSA grants (separated due to their near-exclusive focus on service provision), a different pattern emerged, with California’s 6^th^ district (inclusive of northern Sacramento and its surrounding suburbs) and Texas’ 28^th^ district (inclusive of eastern San Antonio down to the U.S.–Mexico border) experiencing the most pronounced losses. Also notable was the broader distribution of harder-hit congressional districts across the country: most were located throughout the Mid-Atlantic, Midwest, Southeast, and Southwest, with comparatively fewer located in the academic and research institution-heavy Northeast. A table of funding lost in each congressional district is provided in Table 1 in the Supplement.

**Figure 1.**
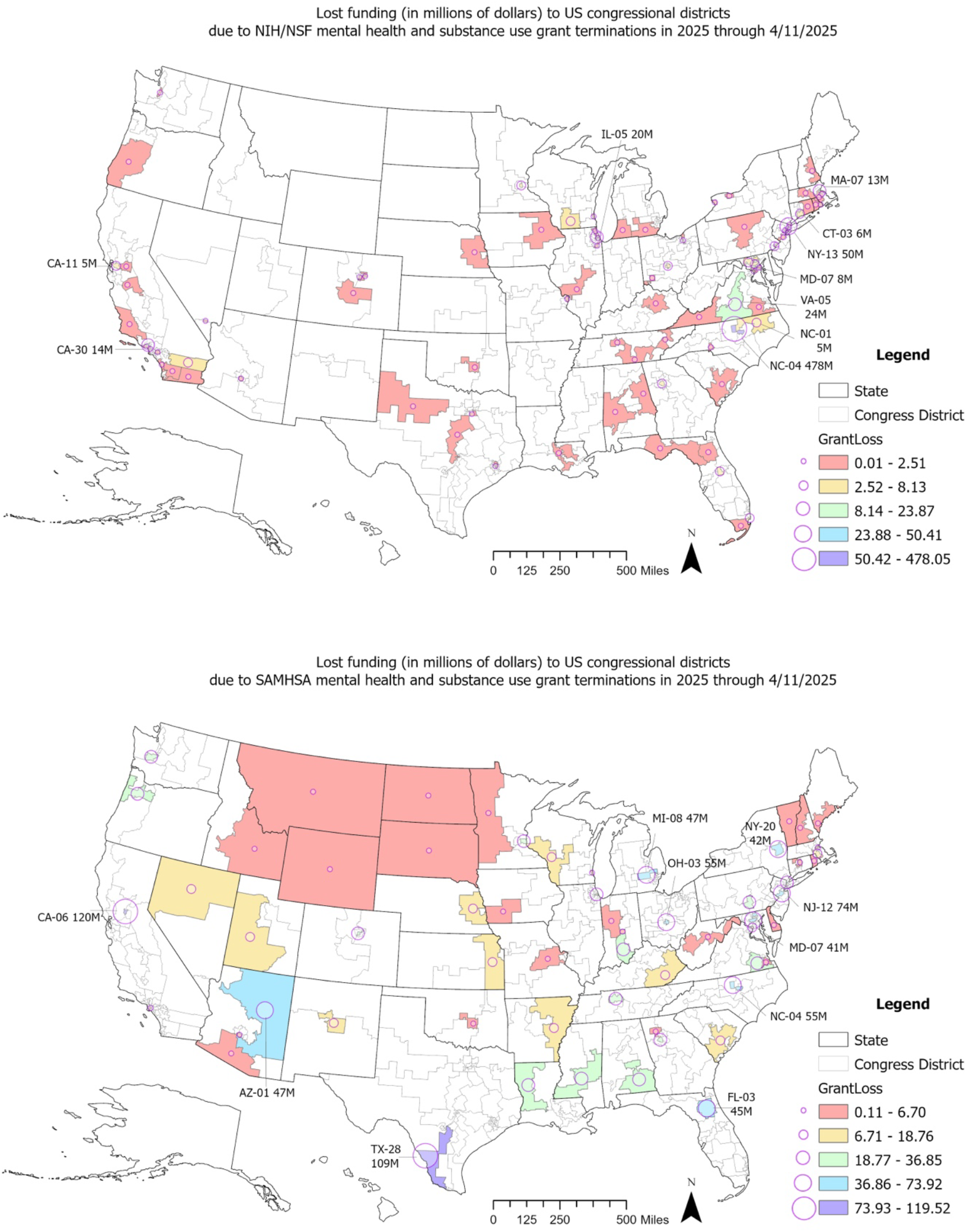
Lost funding (in millions of dollars) to US congressional districts due to NIH/NSF (a, top) and SAMHSA (b, bottom) mental health and substance use grant terminations in 2025 through 4/11/2025

## Discussion

Against a backdrop of ongoing and intersecting mental health and substance use crises, the Trump Administration has slashed research dollars on these topics, creating a chilling effect on the field. We found that nearly $2.1 billion in funding has been lost as of April 25, 2025, including NIH and NSF grants on topics ranging from basic science to equity-related issues and SAMHSA block grants that are used to pay for community mental health and substance use services. Concerningly, the hardest-hit congressional districts were located throughout the Mid-Atlantic, Midwest, Southeast, and Southwest – regions that have been disproportionately burdened by rising rates of depression, suicide, and opioid use, as well as stark socioeconomic inequities that already functioned as significant barriers to mental health and substance use service provision and access.^16–18^ Indeed, aggregating our findings up to the state-level results in a list of hard-hit states that largely parallel those with high rates of suicide and/or substance use deaths (e.g., Arizona, Louisiana, Ohio; see Table 2 in the Supplement),^7,8^ underscoring the potentially deadly consequences of these grant terminations.

These findings extend recent analyses published in *JAMA, Nature*, and *STAT*, and by the U.S. Senate HELP Committee that have quantified the devastating impacts of the Trump Administration’s attacks on science and healthcare. Specifically, such reports have revealed the concerning magnitude by which scientific, medical, and public health research has been cut (upwards of $13.5 billion across HHS);^10–13^ in our study, we used a novel and comprehensive dataset to show that the mental health and substance use fields have likewise been defunded, with considerable geographic variation in which congressional districts lost the most funding. We posit that the concentration of terminations in certain regions likely reflect a confluence of factors, including a disproportionate need for mental health- and substance use-related services (which may lead to a greater emphasis on these topics in their research initiatives), as well as the specific targeting by the NIH and NSF of local institutions such as Columbia University during our study period. Regardless of the cause, however, such cuts are likely to destabilize existing mental health and substance use services and exacerbate inequities between and within U.S. states, ultimately intensifying the challenges faced by local communities (as well as neighboring communities, due to likely spillover effects).^19^

Considering the nature of the terminated grants further reveals the gravity of the situation. In addition to the SAMHSA block grants that provide funds for essential resources such as “priority treatment and support services for individuals without insurance” and “universal, selective, and indicated prevention activities,”^20^ terminated NIH and NSF grants covered a range of topics and methodologies representing the cutting edge of mental health and substance use research. For example, one terminated NIH grant focused on elucidating the neurodevelopmental basis of obsessive-compulsive disorder among youth and proposed using machine learning approaches to identify intervention targets. Another sought to answer the question as to whether maternal COVID-19 infections during pregnancy will have long-term neurodevelopmental effects on children who were born during the pandemic (and if so, how to best intervene). There will be myriad negative consequences of these and the rest of the cuts, including: (1) impaired progress of our understanding of mental health and substance use etiology, treatment, and prevention; (2) halted trials and resultant patient harms; (3) the loss of irreplaceable research data, resulting in a vast waste of taxpayer dollars; and (4) the entrenchment and/or widening of mental health and substance use inequities affecting sexual and gender minorities, people of color, poor and low-income people, people living with disabilities, and people living in rural communities, due to targeted terminations of research into their drivers. More broadly, these terminations are likely to have an enormous toll on the communities surrounding research institutions, which contribute substantially to local economies. Further, given the high number of training grants that were terminated, they could lead to the loss of a generation or more of promising mental health and substance use scientists.

We implore policymakers, policy advocates, and fellow scientists to use our findings to help resist and overturn these unprecedented threats to the mental health and substance use fields, and science, medicine, and public health more generally. Policymakers are in a unique position to effect change and can use our findings as evidence to bolster calls for HHS officials to reinstate funding for terminated grants, support legislation that would shield scientific research from political interference, and conduct oversight hearings. Policy advocates can use our maps to raise awareness of the scientific and economic impacts of mental health- and substance use-related grant terminations on their local communities, such as by organizing town hall meetings, hosting online forums, and speaking with local media. And finally, fellow scientists may use our findings to advocate for change as community members and constituents themselves; further, they can apply our methods to other pertinent topic areas (e.g., cancer, reproductive health) to help reveal the full breadth of harms enacted by the Trump Administration. Ultimately, we hope that this work will support urgently needed efforts aimed at resisting the ongoing political interference and the decimation of the American scientific enterprise.

## Data Availability

Data used in this study are publicly available at: https://grant-witness.us/

## Supplementary Materials

### Search strategy

#### Mental health keywords

“ADHD”, “anorexia nervosa”, “anorexic”, “anxiety”, “ARFID”, “attention-deficit/hyperactivity disorder”, “avoidant restrictive food intake disorder”, “binge eating disorder”, “bipolar disorder”, “borderline personality disorder”, “bulimia nervosa”, “bulimic”, “crisis intervention”, “depression”, “disordered eating”, “disordered weight control behaviors”, “eating disorder”, “emotional well-being”, “mental disorders”, “mental health”, “mental health services”, “mental illness”, “mental well-being”, “neurodevelopmental disorders”, “OSFED”, “other specified feeding or eating disorder”, “personality disorder”, “pica”, “post-traumatic stress disorder”, “psychiatric”, “psychiatric comorbidity”, “psychological distress”, “psychopathology”, “psychosis”, “PTSD”, “purging disorder”, “schizophrenia”, “serious mental illness”, “severe mental illness”, “tele-mental health”, “telepsychiatry”, “telemental health”, and/or “trauma”

#### Substance use keywords

“addiction”, “alcohol abuse”, “alcohol dependence”, “alcohol use”, “alcohol use disorder”, “behavioral health crisis”, “buprenorphine”, “cannabis use”, “cigarette use”, “club drugs”, “codeine”, “cocaine use”, “crisis hotline”, “crisis intervention”, “drug abuse”, “drug addiction”, “drug testing”, “drug use”, “e-cigarettes”, “ecstasy”, “fentanyl”, “GHB”, “hallucinogen use”, “harm reduction”, “heroin”, “hydrocodone”, “illicit drug use”, “injection drug use”, “intoxication”, “MDMA”, “marijuana use”, “medication-assisted treatment”, “mental health crisis”, “mental health emergency”, “methadone”, “methamphetamine use”, “morphine”, “Naloxone”, “Narcan”, “new psychoactive substances”, “nicotine addiction”, “non-suicidal self-injury”, “NSSI”, “opioid”, “opioid use disorder”, “oxycodone”, “OxyContin”, “overdose”, “Percocet”, “prescription drug abuse”, “relapse prevention”, “sedative use”, “self-harm”, “self-injury”, “stimulant use”, “Suboxone Subutex”, “substance abuse”, “substance misuse”, “substance use”, “substance use disorder”, “suicidal”, “suicidality”, “suicide”, “Suicide & Crisis Lifeline”, “synthetic drugs”, “tobacco use”, “tramadol”, “tranquilizer use”, “vape”, “vaping”, “Vicodin Norco”, “withdrawal”, “988 crisis line”, and/or “988 hotline”

---

**Supplementary Table 1.** Funding lost due to terminated mental health- and substance use-related NIH, NSF, and SAMHSA grants by congressional district.

| State | District | Total funding lost | NIH and NSF | SAMHSA |
| --- | --- | --- | --- | --- |
| AL | AL-02 | 22575307.69 | 0.00 | 22575307.69 |
| AL | AL-03 | 872392.11 | 872392.11 | 0.00 |
| AL | AL-07 | 600378.06 | 600378.06 | 0.00 |
| AR | AR-01 | 11924642.91 | 0.00 | 11924642.91 |
| AZ | AZ-01 | 46859362.21 | 0.00 | 46859362.21 |
| AZ | AZ-03 | 1022052.09 | 0.00 | 1022052.09 |
| AZ | AZ-07 | 578011.57 | 471528.35 | 106483.22 |
| CA | CA-06 | 119518402.90 | 0.00 | 119518402.90 |
| CA | CA-44 | 314308.46 | 0.00 | 314308.46 |
| CA | CA-30 | 13510250.63 | 13510250.63 | 0.00 |
| CA | CA-11 | 5366134.07 | 5366134.07 | 0.00 |
| CA | CA-36 | 4464456.40 | 4464456.40 | 0.00 |
| CA | CA-49 | 1921246.00 | 1921246.00 | 0.00 |
| CA | CA-16 | 1920779.78 | 1920779.78 | 0.00 |
| CA | CA-51 | 1002495.22 | 1002495.22 | 0.00 |
| CA | CA-24 | 454773.67 | 454773.67 | 0.00 |
| CA | CA-09 | 449144.23 | 449144.23 | 0.00 |
| CA | CA-33 | 437384.32 | 437384.32 | 0.00 |
| CA | CA-39 | 356728.18 | 356728.18 | 0.00 |
| CA | CA-37 | 245937.27 | 245937.27 | 0.00 |
| CA | CA-50 | 192277.78 | 192277.78 | 0.00 |
| CA | CA-43 | 0.00 | 0.00 | 0.00 |
| CO | CO-01 | 32655749.17 | 1080021.09 | 31575728.08 |
| CO | CO-06 | 925389.22 | 925389.22 | 0.00 |
| CO | CO-05 | 495877.37 | 495877.37 | 0.00 |
| CT | CT-01 | 6115359.16 | 0.00 | 6115359.16 |
| CT | CT-03 | 5897532.26 | 5897532.26 | 0.00 |
| CT | CT-02 | 1083095.51 | 1083095.51 | 0.00 |
| DC | DC-98 | 11323319.10 | 951506.53 | 10371812.57 |
| DC | DC-00 | 3802591.92 | 3802591.92 | 0.00 |
| DE | DE-00 | 2592493.80 | 0.00 | 2592493.80 |
| FL | FL-03 | 46118374.55 | 838100.36 | 45280274.19 |
| FL | FL-27 | 3790245.00 | 3790245.00 | 0.00 |
| FL | FL-10 | 3209158.47 | 3209158.47 | 0.00 |
| FL | FL-26 | 991311.00 | 991311.00 | 0.00 |
| FL | FL-02 | 912375.47 | 912375.47 | 0.00 |
| GA | GA-05 | 41036089.58 | 4190143.15 | 36845946.43 |
| GA | GA-11 | 455703.05 | 0.00 | 455703.05 |
| IA | IA-03 | 6067090.34 | 0.00 | 6067090.34 |
| IA | IA-01 | 487399.73 | 487399.73 | 0.00 |
| ID | ID-02 | 6703503.36 | 0.00 | 6703503.36 |
| IL | IL-07 | 29545908.34 | 747142.34 | 28798766.00 |
| IL | IL-05 | 20356556.23 | 20356556.23 | 0.00 |
| IL | IL-01 | 1697446.70 | 1697446.70 | 0.00 |
| IL | IL-09 | 1553089.62 | 1553089.62 | 0.00 |
| IL | IL-10 | 550599.44 | 550599.44 | 0.00 |
| IL | IL-13 | 88997.21 | 88997.21 | 0.00 |
| IN | IN-09 | 26355892.32 | 0.00 | 26355892.32 |
| IN | IN-07 | 4741123.27 | 0.00 | 4741123.27 |
| IN | IN-04 | 366823.00 | 0.00 | 366823.00 |
| KS | KS-02 | 9617907.16 | 0.00 | 9617907.16 |
| KY | KY-05 | 15630888.84 | 0.00 | 15630888.84 |
| KY | KY-06 | 141024.43 | 141024.43 | 0.00 |
| LA | LA-04 | 34535357.16 | 0.00 | 34535357.16 |
| LA | LA-06 | 417730.00 | 417730.00 | 0.00 |
| MA | MA-04 | 10414632.38 | 840344.39 | 9574287.99 |
| MA | MA-07 | 18781978.46 | 12689266.16 | 6092712.30 |
| MA | MA-08 | 1435338.99 | 1435338.99 | 0.00 |
| MA | MA-02 | 99974.00 | 99974.00 | 0.00 |
| MD | MD-07 | 49253062.71 | 8133497.05 | 41119565.66 |
| MD | MD-08 | 4000000.00 | 4000000.00 | 0.00 |
| MD | MD-04 | 1124844.28 | 1124844.28 | 0.00 |
| ME | ME-01 | 2966950.94 | 0.00 | 2966950.94 |
| MI | MI-08 | 47441780.98 | 0.00 | 47441780.98 |
| MI | MI-06 | 2065186.62 | 2065186.62 | 0.00 |
| MI | MI-07 | 678235.74 | 678235.74 | 0.00 |
| MN | MN-04 | 27492429.61 | 0.00 | 27492429.61 |
| MN | MN-07 | 602982.70 | 0.00 | 602982.70 |
| MN | MN-05 | 4476673.27 | 4476673.27 | 0.00 |
| MO | MO-03 | 2524147.60 | 0.00 | 2524147.60 |
| MO | MO-01 | 684030.55 | 684030.55 | 0.00 |
| MS | MS-03 | 22906208.46 | 0.00 | 22906208.46 |
| MT | MT-00 | 2042892.28 | 0.00 | 2042892.28 |
| MT | MT-01 | 70780.00 | 70780.00 | 0.00 |
| NC | NC-04 | 532688224.40 | 478046501.30 | 54641723.06 |
| NC | NC-01 | 4609153.79 | 4609153.79 | 0.00 |
| NC | NC-12 | 804112.14 | 804112.14 | 0.00 |
| ND | ND-00 | 6324236.00 | 0.00 | 6324236.00 |
| NE | NE-01 | 19925829.91 | 1168090.01 | 18757739.90 |
| NH | NH-02 | 4417554.05 | 0.00 | 4417554.05 |
| NH | NH-01 | 242710.76 | 242710.76 | 0.00 |
| NJ | NJ-12 | 74099759.08 | 181873.00 | 73917886.08 |
| NJ | NJ-10 | 2340568.39 | 2340568.39 | 0.00 |
| NM | NM-01 | 9786434.15 | 0.00 | 9786434.15 |
| NV | NV-02 | 13012827.61 | 0.00 | 13012827.61 |
| NV | NV-01 | 366347.30 | 366347.30 | 0.00 |
| NY | NY-20 | 41911308.67 | 0.00 | 41911308.67 |
| NY | NY-13 | 77699262.89 | 50414601.40 | 27284661.49 |
| NY | NY-26 | 827673.93 | 827673.93 | 0.00 |
| NY | NY-10 | 448474.91 | 448474.91 | 0.00 |
| NY | NY-25 | 375687.63 | 375687.63 | 0.00 |
| NY | NY-12 | 323288.15 | 323288.15 | 0.00 |
| NY | NY-16 | 114680.93 | 114680.93 | 0.00 |
| NY | NY-15 | 48974.00 | 48974.00 | 0.00 |
| OH | OH-03 | 57581979.30 | 2782469.28 | 54799510.02 |
| OH | OH-01 | 1491006.66 | 1491006.66 | 0.00 |
| OH | OH-11 | 279366.00 | 279366.00 | 0.00 |
| OK | OK-05 | 5085590.57 | 514065.66 | 4571524.91 |
| OR | OR-05 | 21893352.36 | 0.00 | 21893352.36 |
| OR | OR-04 | 1224995.89 | 1224995.89 | 0.00 |
| PA | PA-10 | 31164923.96 | 0.00 | 31164923.96 |
| PA | PA-03 | 3322391.39 | 3322391.39 | 0.00 |
| PA | PA-12 | 2138038.14 | 2138038.14 | 0.00 |
| PA | PA-02 | 243811.07 | 243811.07 | 0.00 |
| RI | RI-02 | 4081682.25 | 1418277.16 | 2663405.09 |
| RI | RI-01 | 1092088.27 | 1092088.27 | 0.00 |
| SC | SC-06 | 13461993.91 | 450812.87 | 13011181.04 |
| SD | SD-00 | 2813280.96 | 0.00 | 2813280.96 |
| TN | TN-05 | 27879764.10 | 1297138.07 | 26582626.03 |
| TN | TN-02 | 32901.29 | 32901.29 | 0.00 |
| TN | TN-04 | 14563.12 | 14563.12 | 0.00 |
| TX | TX-28 | 109427851.20 | 0.00 | 109427851.20 |
| TX | TX-25 | 1336740.25 | 1336740.25 | 0.00 |
| TX | TX-30 | 900819.12 | 900819.12 | 0.00 |
| TX | TX-19 | 216855.09 | 216855.09 | 0.00 |
| TX | TX-18 | 36978.00 | 36978.00 | 0.00 |
| UT | UT-02 | 8990798.36 | 0.00 | 8990798.36 |
| VA | VA-04 | 34038451.48 | 15554.59 | 34022896.89 |
| VA | VA-03 | 2790936.96 | 0.00 | 2790936.96 |
| VA | VA-05 | 23870599.92 | 23870599.92 | 0.00 |
| VA | VA-09 | 1275364.74 | 1275364.74 | 0.00 |
| VT | VT-00 | 1672282.84 | 0.00 | 1672282.84 |
| WA | WA-10 | 32591064.68 | 0.00 | 32591064.68 |
| WA | WA-07 | 1191921.40 | 1191921.40 | 0.00 |
| WI | WI-03 | 16746506.10 | 0.00 | 16746506.10 |
| WI | WI-04 | 5473632.28 | 2511689.48 | 2961942.80 |
| WI | WI-02 | 2693639.80 | 2693639.80 | 0.00 |
| WV | WV-02 | 6559806.29 | 0.00 | 6559806.29 |
| WY | WY-00 | 1664120.80 | 0.00 | 1664120.80 |

**Supplementary Table 2.**
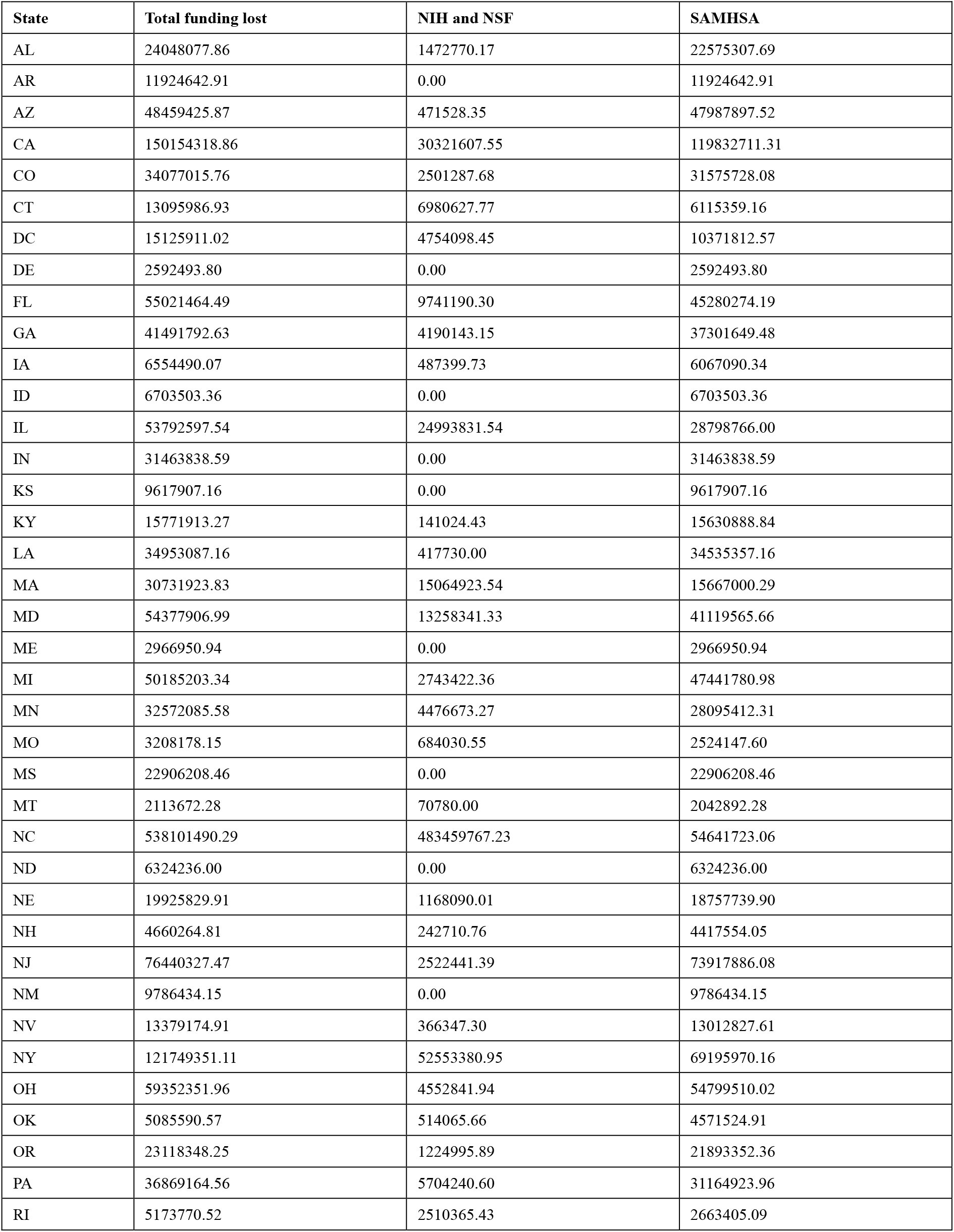
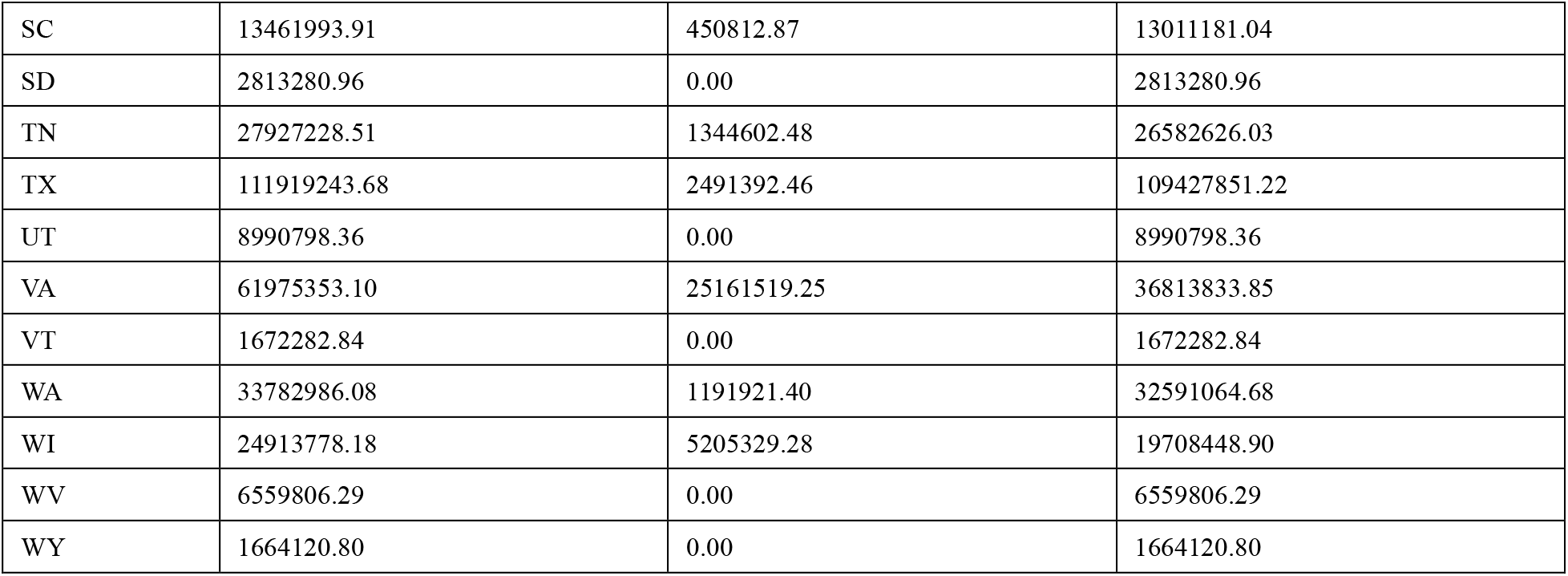
Funding lost due to terminated mental health- and substance use-related NIH, NSF, and SAMHSA grants by state.

| State | Total funding lost | NIH and NSF | SAMHSA |
| --- | --- | --- | --- |
| AL | 24048077.86 | 1472770.17 | 22575307.69 |
| AR | 11924642.91 | 0.00 | 11924642.91 |
| AZ | 48459425.87 | 471528.35 | 47987897.52 |
| CA | 150154318.86 | 30321607.55 | 119832711.31 |
| CO | 34077015.76 | 2501287.68 | 31575728.08 |
| CT | 13095986.93 | 6980627.77 | 6115359.16 |
| DC | 15125911.02 | 4754098.45 | 10371812.57 |
| DE | 2592493.80 | 0.00 | 2592493.80 |
| FL | 55021464.49 | 9741190.30 | 45280274.19 |
| GA | 41491792.63 | 4190143.15 | 37301649.48 |
| IA | 6554490.07 | 487399.73 | 6067090.34 |
| ID | 6703503.36 | 0.00 | 6703503.36 |
| IL | 53792597.54 | 24993831.54 | 28798766.00 |
| IN | 31463838.59 | 0.00 | 31463838.59 |
| KS | 9617907.16 | 0.00 | 9617907.16 |
| KY | 15771913.27 | 141024.43 | 15630888.84 |
| LA | 34953087.16 | 417730.00 | 34535357.16 |
| MA | 30731923.83 | 15064923.54 | 15667000.29 |
| MD | 54377906.99 | 13258341.33 | 41119565.66 |
| ME | 2966950.94 | 0.00 | 2966950.94 |
| MI | 50185203.34 | 2743422.36 | 47441780.98 |
| MN | 32572085.58 | 4476673.27 | 28095412.31 |
| MO | 3208178.15 | 684030.55 | 2524147.60 |
| MS | 22906208.46 | 0.00 | 22906208.46 |
| MT | 2113672.28 | 70780.00 | 2042892.28 |
| NC | 538101490.29 | 483459767.23 | 54641723.06 |
| ND | 6324236.00 | 0.00 | 6324236.00 |
| NE | 19925829.91 | 1168090.01 | 18757739.90 |
| NH | 4660264.81 | 242710.76 | 4417554.05 |
| NJ | 76440327.47 | 2522441.39 | 73917886.08 |
| NM | 9786434.15 | 0.00 | 9786434.15 |
| NV | 13379174.91 | 366347.30 | 13012827.61 |
| NY | 121749351.11 | 52553380.95 | 69195970.16 |
| OH | 59352351.96 | 4552841.94 | 54799510.02 |
| OK | 5085590.57 | 514065.66 | 4571524.91 |
| OR | 23118348.25 | 1224995.89 | 21893352.36 |
| PA | 36869164.56 | 5704240.60 | 31164923.96 |
| RI | 5173770.52 | 2510365.43 | 2663405.09 |
| SC | 13461993.91 | 450812.87 | 13011181.04 |
| SD | 2813280.96 | 0.00 | 2813280.96 |
| TN | 27927228.51 | 1344602.48 | 26582626.03 |
| TX | 111919243.68 | 2491392.46 | 109427851.22 |
| UT | 8990798.36 | 0.00 | 8990798.36 |
| VA | 61975353.10 | 25161519.25 | 36813833.85 |
| VT | 1672282.84 | 0.00 | 1672282.84 |
| WA | 33782986.08 | 1191921.40 | 32591064.68 |
| WI | 24913778.18 | 5205329.28 | 19708448.90 |
| WV | 6559806.29 | 0.00 | 6559806.29 |
| WY | 1664120.80 | 0.00 | 1664120.80 |

## Notes

### Competing Interest Statement

The authors have declared no competing interest.

### Funding Statement

This study did not receive any funding.

